# *In vivo* virological efficacy of monoclonal antibodies and direct antiviral agents against the SARS-CoV-2 BA.1 and BA.2 Omicron sublineages

**DOI:** 10.1101/2022.06.23.22276509

**Authors:** V Mazzotta, A Cozzi Lepri, F Colavita, S Rosati, E Lalle, C Cimaglia, J Paulicelli, I Mastrorosa, A Vergori, E Girardi, AR Garbuglia, F Vaia, E Nicastri, A Antinori

**Author notes:** **Corresponding author** Valentina Mazzotta, MD, Clinical and Research Infectious Diseases Department, National Institute for Infectious Diseases Lazzaro Spallanzani IRCCS, Via Portuense 292, 00149 Roma, Italy.

## Abstract

**Background:** Omicron variant questioned the efficacy of the approved therapies for the early COVID-19. *I In vitro* data show retained neutralizing activity against BA.1 and BA.2 for remdesivir (RDV), molnupiravir (MLN), and nirmatrelvir/ritonavir (NRM/r), while poor efficacy for Sotrovimab (STR) against BA.2. No data about the risk of clinical failure and *in vivo* antiviral activity are available.

**Material and methods:** Single-center observational comparison study enrolling all consecutive patients with a confirmed SARS-CoV-2 Omicron (BA.1 or BA.2) diagnosis and who met eligibility criteria for treatment with RDV, MLN, NRM/r, or STR. Treatment allocation was subject to drug availability, time from symptoms onset, and comorbidities. Patients were followed through day 30. Nasopharyngeal swab (NPS) VL was measured on day 1 (D1) and D7 and was expressed by log_2_ cycle threshold (CT) scale. Comparisons between groups were made by Chi-square and Wilcoxon paired-test. Primary endpoint was D1-D7 VL variation. Potential decrease in VL and average treatment effect (ATE) were calculated from fitting marginal linear regression models weighted for calendar month of infusion, duration of symptoms, and immunodeficiency. Secondary endpoints were the proportion of D7 undetectable VL in NPS and clinical outcomes compared by treatment groups using a Chi-square test.

**Results:** A total of 521 pts received treatments (STR 202, MLN 117, NRM/r 84, and RDV 118): female 250 (48%), median age 66 yrs (IQR 55-76), 90% vaccinated; 15% with negative baseline serology. At D1, median time from symptoms onset was 3 days (2,4). 378 (73%) pts were infected with BA.1 and 143 (27%) with BA.2. D1 mean viral load was 4.12 log2 (4.16 for BA.1 and 4.01 for BA.2). The adjusted analysis showed that NRM/r significantly reduced VL compared to all the other drugs in pts infected with BA.1 while no evidence for a difference vs. MLP was seen in those infected with BA.2. MLN had comparable activity to STR against BA.1 and to NRM/r against BA.2. There was no significant difference between STR and RDV for BA.2.

At D7, 35/521 (6.7%) pts had undetectable VL. Of these, 31 were infected with BA.1 [9 (9%) MLN, 7 (14%) NRM/r, 7 (8%) RDV, and 8 (5%) STR)], and only 4 with BA.2, all treated with NRM/r. After 30 days of follow-up, 9/568 pts experienced COVID-19-related clinical failure [7/226 STR (5 BA.1) and 2/87 NRM /r (2 BA.1)].

**Conclusions:** In this analysis of *in vivo* early VL reductions, NRM/r appears to be the drug showing the greatest antiviral activity regardless of the VoC, together with MLN, although the latter limited to people with BA.2. In the Omicron era, due to the high prevalence of vaccinated people and the lower probability of hospital admission, VL decrease can be a valuable surrogate of drug activity.

## MAIN TEXT

The Omicron (B.1.1.529) variant of severe acute respiratory syndrome coronavirus 2 (SARS-CoV-2), and its sublineages BA.1 and BA.2, have become the predominant variants responsible for coronavirus disease 2019 (COVID-19) circulating worldwide. The large number of critical mutations in Spike protein of these subvariants raised concerns about the efficacy of therapies for the early phase of COVID-19, particularly of monoclonal antibodies (mAbs).

Previously published *in vitro* data showed that mAbs combination Bamlanivimab/Etesevimab and Casirivimab/Imdevimab showed little neutralizing activity against BA.1 and BA.2; conversely, Sotrovimab retained most of activity against omicron/BA.1, but was escaped by omicron/BA.2, with a 16 to 37 fold-reduction in neutralizing activity; finally, Tixagevimab/Cilgavimab retained most of activity against BA.2, but it was not as effective against BA.1^1–2^. Differently from mAbs, antiviral agents, such as Remdesivir, Molnupiravir, or Nirmatrelvir/ritonavir, which target the highly conserved protein of SARS-CoV-2, consistently retained *in vitro* activity against both BA.1 and BA.2 sublineages^3–4^.

Analyses of *in vivo* data evaluating the clinical efficacy of these agents against the new variant are lacking. Primary endpoint in phase-3 randomized studies in COVID-19 was typically the proportion of participants hospitalized or dead after randomization. Due to the lower risk of severe outcomes following SARS-CoV-2 Omicron infection^5^, and taking into account the high prevalence of vaccinated people during the Omicron wave, a clinical outcome is not suited to the current scenario. Viral load reduction from baseline through day 7 was used as endpoint of phase-2 studies of mAbs and may be a valuable surrogate marker of *in vivo* neutralizing or antiviral activity.

We assessed the *in vivo* viral load reduction in nasopharyngeal swab collected on day1 and day7 from outpatients treated with Sotrovimab, Molnupiravir, Remdesivir, or Nirmatrelvir/ritonavir for mild-to-moderate COVID-19 due to sublineages BA.1 or BA.2.

Details on patient recruitment, inclusion criteria, testing, and statistical analyses are included in the Supplements.

Of 568 participants enrolled, 521 had a viral load measured at day7: 202 received Sotrovimab, 117 Molnupiravir, 84 Nirmatrelvir/ritonavir and 118 Remdesivir. Overall, 250 (48%) were female, 469 (90%) were vaccinated and 81 (15%) had negative baseline serology. Median age was 66 years (IQR 55-76) and median time from symptoms onset to day1 was 3 days (2-4). BA.1 and BA.2 were detected in 378 (73%) and 143 (27%), respectively. A higher proportion of chronic respiratory disease (chi-square, p<0·001), liver disease (p<0·001) and immunodeficiency (p=0·01) was observed at day1 among participants receiving Sotrovimab. Baseline mean viral load was 4·12 (SD 0·27) log_2_ CT [4·16 for BA.1 and 4·01 for BA.2]. Detailed characteristics according to treatment groups are reported in Supplementary Table 1. Linear regression analysis calculating the average treatment effect of therapies when compared to each other in separately emulated parallel trials showed that Nirmatrelvir/ritonavir significantly reduced viral load compared to other drugs both in the BA.1 and BA.2 subgroups (Figure 1a and 1b). In contrast, there was no difference in activity against BA.1 between Sotrovimab and Molnupiravir and between the latter and Nirmatrelvir/ritonavir against BA.2. There was no evidence for a difference in activity between Sotrovimab and Remdesivir against both the two subvariants. All variations of SARS-CoV-2 RNA levels from day1 to day7 according to treatment groups are reported in Supplementary Figure 1. Detailed results of potential decrease in viral load and average treatment effect for all possible 2-by-2 treatment comparisons separately for BA.1 and BA.2 are also shown in Supplementary tables 2 and 3. Proportion of participants with CT≤40 at day7 was 6·7% (35/521, 31 infected with BA.1 and 4 with BA.2). See details in Supplementary table 4.

**Figure 1.**
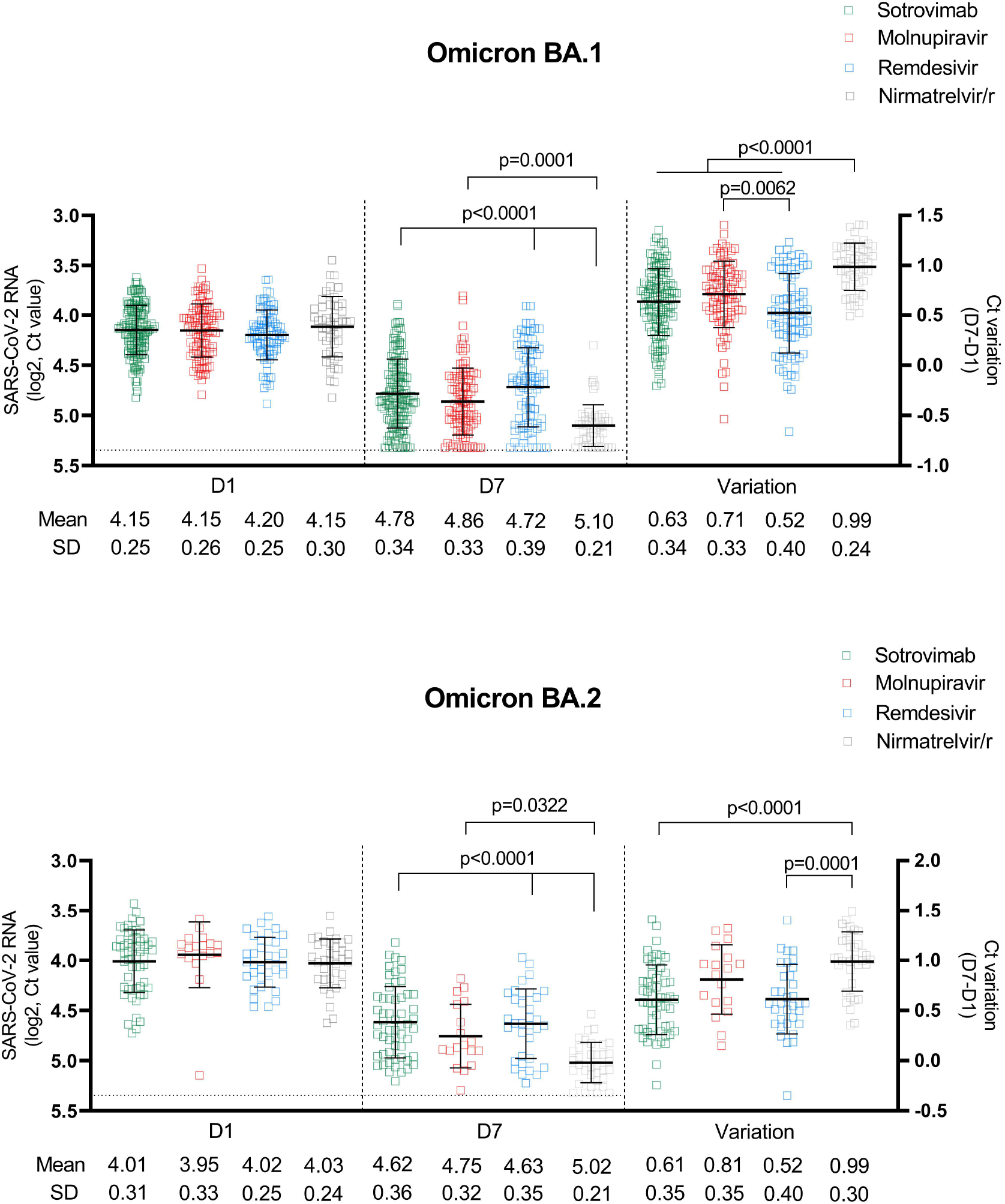
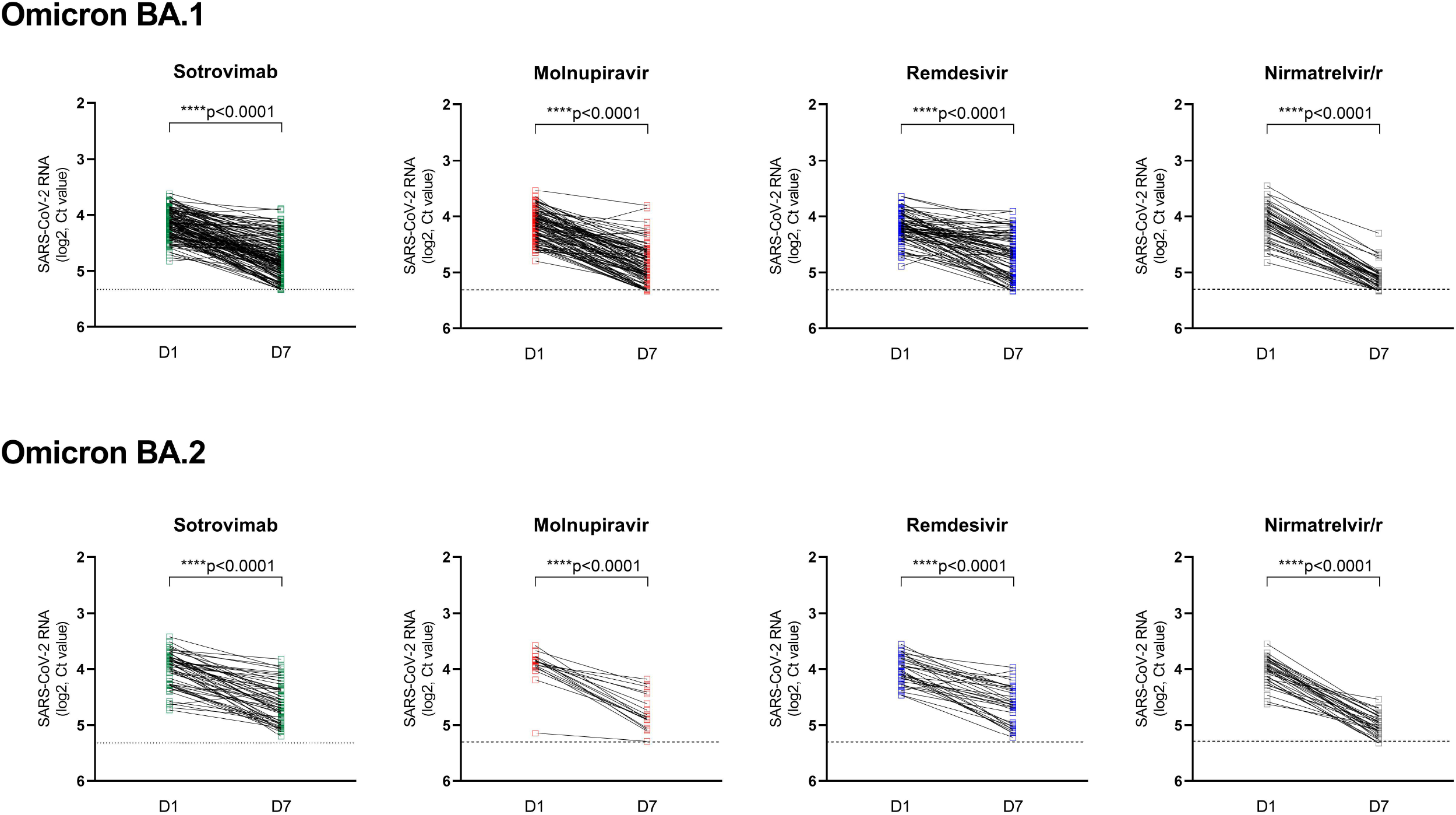
SARS-CoV-2 RNA levels at D1 and D7 in patients treated with Sotrovimab, Molnupiravir, Remdesivir and Nirmatrelvir/r. Dot-plots showing the comparison of viral loads detected at D1 and D7 and the variation of RNA levels observed between the two time-points by intervention in **(A)** patients with Omicron BA.1 infection treated with Sotrovimab (n=146), or Molnupiravir (n=99), or Remdesivir (n=84), or Nirmatrelvir/r (n=49); **(B)** patients with Omicron BA.2 infection treated with Sotrovimab (n=56), or Molnupiravir (n=18), or Remdesivir (n=34), or Nirmatrelvir/r (n=35). Viral RNA levels are expressed as log2 CT values. Mean of log2 CT values and SD are shown. Statistical analysis of the comparisons between treatment groups was performed by Kruskal-Wallis test, adjusted with Dunn’s multiple comparisons test. Horizontal dashed line represents the limit of detection (CT: 40.0), values ≥40 are considered negative.

COVID-19-related hospitalization or death from any cause through day 30 was assessed in 568 patients: 9 patients [7/226 (3·1%) Sotrovimab (5 BA.1) and 2/87 (2·3%) Nirmatrelvir/ritonavir (2 BA.1)] experienced clinical failure.

The main limitations of our analysis are the observational nature of the study conducted in a single COVID-19 health care center and the lack of a randomized design, which does not allow to rule out confounding bias. These limitations are partially mitigated by the use of weighted marginal linear regression models and appropriate control of measured confounding factors. Our results are however important as, to the best of our knowledge, this is the first analysis to evaluate the *in vivo* efficacy of currently available treatments against the Omicron variants.

In conclusion, according to our viral load change dynamic model and assumptions, in outpatients with mild-to-moderate COVID-19, Nirmatrelvir/ritonavir appears to be the option with the strongest *in vivo* antiviral activity against Omicron variant among all other treatment options examined. Only for Molnupiravir and limited to the BA.2 sublineage, antiviral effect appeared to be comparable to that observed with Nirmatrelvir/ritonavir. Because of the low incidence of hospital admissions in the Omicron era, emulation of trials with surrogate endpoints such as *in vivo* neutralizing activity can provide useful information for treatment decisions of early COVID-19.

## Data Availability

All data produced in the present study are available upon reasonable request to the authors

## Data sharing

Anonymized participant data will be made available upon reasonable requests directed to the corresponding author. Proposals will be reviewed and approved by investigator, and collaborators on the basis of scientific merit. After approval of a proposal, data can be shared through a secure online platform after signing a data access agreement.

## Acknowledgments

The authors gratefully acknowledge the nurse staff and all the patients.

