## Supplementary materials for "*In vivo* virological efficacy of monoclonal antibodies and direct antiviral agents against the SARS-CoV-2 BA.1 and BA.2 Omicron sublineages"

Mazzotta Valentina, Cozzi Lepri Alessandro, Colavita Francesca, Rosati Silvia, Lalle Eleonora, Cimaglia Claudia, Paulicelli Jessica, Mastrorosa Ilaria, Vergori Alessandra, Girardi Enrico, Garbuglia Anna Rosa, Vaia Francesco, Nicastri Emanuele, Antinori Andrea.

**Supplementary Methods**

This analysis uses the data of an observational study on the effectiveness of early treatment for outpatients with mild-to-moderate COVID-19. The study was approved by the Scientific Committee of the Italian Drug Agency (AIFA) and by the Ethical Committee of the Lazzaro Spallanzani Institute, as National Review Board for COVID-19 pandemic in Italy (approval number 380/2021).

All consecutive patients presenting from 21st of December, 2021 to 15th of March, 2022 to the National Institute for Infectious Diseases “L. Spallanzani” with a confirmed SARS-CoV-2 Omicron (BA.1 or BA.2) diagnosis and a mild-to-moderate COVID-19, who met the Italian Medicine Agency (AIFA) criteria for eligibility for early treatment by mAbs or antiviral agents were enrolled. Treatment allocation was subject to drug availability, time from symptoms onset, and presence of comorbidities as defined by AIFA criteria.

Outpatients visits, with medical evaluation, vital signs recording and laboratory tests, were scheduled at baseline (day of treatment, day1) and at seven day after (day7). Patients were followed-up for occurrence of clinical events through day 30 after starting treatment through a telephone visit.

Viral load in nasopharyngeal swab was assessed using Abbott Alinity m RealTime System (Abbott Laboratories, Wiesbaden, Germany) on day1 (baseline) and day7, and expressed as log2 of cycle threshold (CT) values. Identification of SARS-CoV-2 variants was performed by Sanger sequencing of the Spike coding gene on samples collected on day1. SARS-COV-2 serology was performed by two chemiluminescence microparticle assays (CMIA) detecting anti-Nucleoprotein and anti-Spike/RBD IgG (ARCHITECT SARS-CoV-2 IgG, and ARCHITECT SARS-CoV-2 IgG II Quantitative; Abbott Laboratories, Wiesbaden, Germany, respectively). According to the to manufacturer’s instructions, for the two CMIA, Index >1.4 and Binding Antibody Units (BAU)/mL ≥7.1 are considered positive for anti-N and anti-Spike/RBD IgG, respectively.

Primary endpoint was log2 viral load variation from day1 to day7. We adopted the log transformation because the distribution of the viral load change in the raw scale was positively skewed and significantly deviating from the normal distribution. Secondary endpoints were the proportion of negative nasopharyngeal swab at day7 and the proportion of patients who experienced COVID-related clinical failure, defined as hospitalization due to development of severe COVID-19 or death from any cause over days 0-30. Main characteristics of the participants, assessed at day1, were compared by treatment strategy using Chi-square (categorical variables) and Kruskal-Wallis (continuous variable) test. We estimated potential outcomes and the average treatment effect (ATE) of treatment on viral load change at day7. Because we had 4 drugs to compare this led to 6 possible 2-by-2 comparisons in separate parallel trials. We controlled for confounding by modelling the treatment assignment (via inverse probability of weighting) or the outcome (via regression adjustment) or both (doubly robust methods). The latter provides unbiased estimates for the treatment effect even if one of the models is mis-specified. According to our assumptions we identified the following key confounding factors: calendar month of infusion, immunodeficiency at time of infusion and duration of symptoms. All analyses were controlled for these factors.

Proportion of participants who experienced the secondary endpoints were shown by treatment group and compared using a chi-square test. All analyses were stratified by type of Omicron variant detected (BA.1 vs BA.2).

**Supplementary Table 1. Main characteristics at enrolment by intervention**

|  | **Regimen started** | | | | | |
| --- | --- | --- | --- | --- | --- | --- |
| **Characteristics** | **Sotrovimab** | **Molnupiravir** | **Remdesivir** | **Nirmatrelvir/r** | **p-value^*^** | **Total** |
|  | N= 202 | N= 117 | N= 118 | N= 84 |  | N= 521 |
| ***Gender, n(%)*** |  |  |  |  | 0.454 |  |
| Female | 105 (52.0%) | 52 (44.4%) | 52 (44.1%) | 41 (48.8%) |  | 250 (48.0%) |
| ***Age, years*** |  |  |  |  | 0.092 |  |
| Median (IQR) | 63 (52, 75) | 68 (57, 75) | 70 (57, 78) | 63 (55, 76) |  | 66 (55, 76) |
| Older than 65, n(%) | 93 (46.0%) | 65 (55.6%) | 71 (60.2%) | 36 (42.9%) | 0.028 | 265 (50.9%) |
| ***Days from sypmtoms onset to MAbs infusion*** |  |  |  |  | 0.003 |  |
| Median (IQR) | 3 (2, 5) | 3 (2, 4) | 4 (2, 5) | 3 (2, 4) |  | 3 (2, 4) |
| ***Comorbidities/risk factors, n(%)*** |  |  |  |  |  |  |
| Diabetes | 28 (13.9%) | 24 (20.5%) | 23 (19.5%) | 19 (22.6%) | 0.239 | 94 (18.0%) |
| Obesity (BMI>30) | 93 (46.0%) | 64 (54.7%) | 73 (61.9%) | 53 (63.1%) | 0.012 | 283 (54.3%) |
| CVD | 31 (15.3%) | 27 (23.1%) | 24 (20.3%) | 17 (20.2%) | 0.357 | 99 (19.0%) |
| Chronic respiratory disease | 22 (10.9%) | 2 (1.7%) | 1 (0.8%) | 1 (1.2%) | <.001 | 26 (5.0%) |
| Renal impairment | 7 (3.5%) | 1 (0.9%) | 2 (1.7%) | 0 (0.0%) | 0.174 | 10 (1.9%) |
| Hepatic Disease | 61 (30.2%) | 20 (17.1%) | 21 (17.8%) | 9 (10.7%) | <.001 | 111 (21.3%) |
| Cancer | 32 (15.8%) | 15 (12.8%) | 16 (13.6%) | 17 (20.2%) | 0.485 | 80 (15.4%) |
| Primary/secondary immunodeficiency | 52 (25.7%) | 17 (14.5%) | 18 (15.3%) | 10 (11.9%) | 0.010 | 97 (18.6%) |
| Neurologic disease | 16 (7.9%) | 6 (5.1%) | 3 (2.5%) | 4 (4.8%) | 0.229 | 29 (5.6%) |
| ***Vital signs at baseline*** |  |  |  |  |  |  |
| SpO2, median (IQR) | 98 (97, 99) | 98 (97, 99) | 98 (97, 98) | 98 (97, 99) | 0.173 | 98 (97, 99) |
| Fever (>37.5°C), n(%) | 8 (4.2%) | 0 (0.0%) | 2 (1.8%) | 0 (0.0%) | 0.036 | 10 (2.0%) |
| BMI, median (IQR) | 24.65 (22.20, 28.57) | 25.26 (23.23, 29.40) | 26.19 (23.23, 29.02) | 26.06 (23.44, 29.48) | 0.093 | 25.31 (22.99, 29.05) |
| ***Laboratory values, median (IQR)*** |  |  |  |  |  |  |
| Ferritin, ng/ml | 99.00 (39.00, 181.0) | 89.00 (48.00, 173.0) | 84.00 (44.00, 155.0) | 94.00 (55.00, 145.0) | 0.885 | 91.00 (47.00, 171.0) |
| C-reactive protein, mg/dl | 0.95 (0.32, 2.48) | 0.93 (0.29, 1.91) | 0.88 (0.34, 1.67) | 0.99 (0.37, 1.98) | 0.793 | 0.93 (0.32, 2.09) |
| Lymphocytes, /uL | 1270 (870.0, 1670) | 1460 (1145, 2045) | 1470 (1100, 1790) | 1500 (1120, 1940) | <.001 | 1400 (1010, 1840) |
| ***Baseline SARS-COV-2*** |  |  |  |  |  |  |
| ***Serology, n(%)*** |  |  |  |  | 0.006 |  |
| Anti-N positive | 4 (2.0%) | 3 (2.6%) | 3 (2.5%) | 3 (3.6%) |  | 13 (2.5%) |
| Anti-S Positive | 138 (68.3%) | 90 (76.9%) | 97 (82.2%) | 72 (85.7%) |  | 397 (76.2%) |
| Negative | 43 (21.3%) | 16 (13.7%) | 15 (12.7%) | 7 (8.3%) |  | 81 (15.5%) |
| Unknown | 17 (8.4%) | 8 (6.8%) | 3 (2.5%) | 2 (2.4%) |  | 30 (5.8%) |
| ***Vaccination, n(%)*** |  |  |  |  | 0.185 |  |
| Yes (partly or fully) | 182 (91.0%) | 108 (93.1%) | 101 (85.6%) | 78 (92.9%) |  | 469 (90.5%) |
| ***Vaccine type, n(%)*** |  |  |  |  | 0.463 |  |
| BNT162b2 | 87 (73.7%) | 50 (75.8%) | 38 (74.5%) | 39 (75.0%) |  | 214 (74.6%) |
| mRNA-1273 | 22 (18.6%) | 10 (15.2%) | 10 (19.6%) | 6 (11.5%) |  | 48 (16.7%) |
| ChAdOx1 | 9 (7.6%) | 6 (9.1%) | 3 (5.9%) | 7 (13.5%) |  | 25 (8.7%) |
| Ad26.COV2.S | 0 (0.0%) | 0 (0.0%) | 0 (0.0%) | 0 (0.0%) |  | 0 (0.0%) |
| Other/unknown | 64 (35.2%) | 42 (38.9%) | 50 (49.5%) | 26 (33.3%) |  | 182 (38.8%) |
| ***SARS-COV-2 variant, n(%)*** |  |  |  |  |  |  |
| Omicron BA1 | 146 (72.3%) | 99 (84.6%) | 84 (71.2%) | 49 (58.3%) | <.001 | 378 (72.6%) |
| Omicron BA2 | 56 (27.7%) | 18 (15.4%) | 34 (28.8%) | 35 (41.7%) | <.001 | 143 (27.4%) |
| ***Baseline CT*** |  |  |  |  |  |  |
| *Mean (SD)* | 17.57 ± 3.39 | 17.75 ± 3.72 | 17.99 ± 3.31 | 17.23 ± 3.49 | 0.283 | 17.65 ± 3.46 |
| *log2 scale Mean (SD)* | 4.11 ± 0.27 | 4.12 ± 0.28 | 4.15 ± 0.26 | 4.08 ± 0.28 | 0.283 | 4.12 ± 0.27 |
| *Less than 25, n(%)* | 195 (96.5%) | 114 (97.4%) | 113 (95.8%) | 81 (96.4%) | 0.919 | 503 (96.5%) |
| ***MASS score, median (IQR)*** | 3 (0, 5) | 3 (2, 5) | 3 (2, 5) | 3 (1, 4) | 0.444 | 3 (1, 5) |
| ***Baseline symptoms score, median (IQR)*** | 11 (6, 14) | 11 (7, 16) | 8 (4, 15) | 12 (8, 16) | 0.172 | 10 (6, 15) |
| ^*^Chi-square or Kruskal-Wallis test as appropriate | | | | | | |
| IQR, interquartile range; BMI, body mass index; SpO2, peripheral oxygen saturation; MASS score, Monoclonal Antibody Screening Score. | | | | | | |

**Supplementary table 2. Potential Outcomes and ATE from fitting linear regression models - subset of infected with BA.1**

|  | **Potential Day 7 decrease in CT from baselineand ATE^&^ from fitting a linear regression model (log2 scale)** | | | |
| --- | --- | --- | --- | --- |
|  | **Decrease (log2) in intervention  (95% CI)** | **Decrease (log2) in control  (95% CI)** | **ATE^*^ (95% CI)** | **p-value** |
| ***Sotrovimab vs. Molnupiravir*** |  |  |  |  |
| IPWs | 0.65 (0.59, 0.71) | 0.68 (0.59, 0.77) | -0.03 (-0.14, 0.07) | 0.549 |
| Double Robust | 0.65 (0.59, 0.71) | 0.69 (0.59, 0.78) | -0.04 (-0.15, 0.08) | 0.490 |
| Regression adjustment | 0.65 (0.59, 0.71) | 0.69 (0.59, 0.78) | -0.04 (-0.15, 0.08) | 0.526 |
| Truncated |  |  | -0.07 (-0.22, 0.09) | 0.404 |
| ***Molnupiravir vs. Nirmatrelvir/r*** |  |  |  |  |
| IPWs | 0.68 (0.57, 0.79) | 0.96 (0.88, 1.04) | -0.28 (-0.41, -0.14) | <.001 |
| Double Robust | 0.69 (0.58, 0.79) | 0.95 (0.87, 1.03) | -0.27 (-0.40, -0.13) | <.001 |
| Regression adjustment | 0.68 (0.58, 0.78) | 1.03 (0.95, 1.12) | -0.35 (-0.48, -0.22) | <.001 |
| Truncated |  |  | -0.72 (-1.11, -0.33) | <.001 |
| ***Remdesivir vs. Nirmatrelvir/r*** |  |  |  |  |
| IPWs | 0.48 (0.39, 0.57) | 0.97 (0.89, 1.05) | -0.49 (-0.62, -0.37) | <.001 |
| Double Robust | 0.48 (0.39, 0.58) | 0.93 (0.82, 1.03) | -0.44 (-0.58, -0.30) | <.001 |
| Regression adjustment | 0.48 (0.38, 0.58) | 0.98 (0.88, 1.08) | -0.50 (-0.64, -0.35) | <.001 |
| Truncated |  |  | -1.19 (-1.66, -0.71) | <.001 |
| ***Remdesivir vs. Sotrovimab*** |  |  |  |  |
| IPWs | 0.50 (0.41, 0.60) | 0.66 (0.60, 0.72) | -0.15 (-0.26, -0.04) | 0.007 |
| Double Robust | 0.50 (0.40, 0.60) | 0.66 (0.60, 0.72) | -0.16 (-0.27, -0.04) | 0.006 |
| Regression adjustment | 0.50 (0.41, 0.60) | 0.66 (0.60, 0.72) | -0.15 (-0.27, -0.04) | 0.008 |
| Truncated |  |  | -0.23 (-0.40, -0.06) | 0.008 |
| ***Sotrovimab vs. Nirmatrelvir/r*** |  |  |  |  |
| IPWs | 0.67 (0.61, 0.73) | 1.03 (0.95, 1.10) | -0.36 (-0.46, -0.25) | <.001 |
| Double Robust | 0.67 (0.61, 0.73) | 0.98 (0.90, 1.06) | -0.31 (-0.41, -0.21) | <.001 |
| Regression adjustment | 0.66 (0.60, 0.73) | 1.03 (0.94, 1.12) | -0.37 (-0.48, -0.26) | <.001 |
| Truncated |  |  | -0.63 (-0.93, -0.34) | <.001 |
| ***Molnupiravir vs. Remdesivir*** |  |  |  |  |
| IPWs | 0.69 (0.61, 0.77) | 0.48 (0.38, 0.59) | 0.20 (0.07, 0.34) | 0.003 |
| Double Robust | 0.69 (0.59, 0.78) | 0.49 (0.37, 0.60) | 0.20 (0.05, 0.34) | 0.005 |
| Regression adjustment | 0.69 (0.60, 0.78) | 0.50 (0.40, 0.60) | 0.19 (0.05, 0.32) | 0.006 |
| Truncated |  |  | 0.29 (0.09, 0.49) | 0.005 |
| ^&^Average Treatment Effect | | | | |
| ^*^weighted for month of infusion, duration of symptoms and immunodeficiency | | | | |

**Supplementary table 3. Potential Outcomes and ATE from fitting linear regression models - subset of infected with BA.2**

|  | **Potential Day 7 decrease in CT from baseline and ATE^&^ from fitting a linear regression model (log2 scale)** | | | |
| --- | --- | --- | --- | --- |
|  | **Decrease (log2) in intervention  (95% CI)** | **Decrease (log2) in control  (95% CI)** | **ATE^*^ (95% CI)** | **p-value** |
| ***Sotrovimab vs. Molnupiravir*** |  |  |  |  |
| IPWs | 0.60 (0.51, 0.68) | 0.91 (0.80, 1.02) | -0.31 (-0.45, -0.17) | <.001 |
| Double Robust | 0.60 (0.51, 0.69) | 0.89 (0.72, 1.07) | -0.29 (-0.50, -0.09) | <.001 |
| Regression adjustment | 0.60 (0.51, 0.70) | 0.89 (0.75, 1.04) | -0.29 (-0.46, -0.12) | <.001 |
| Truncated |  |  | -0.25 (-0.47, -0.03) | 0.025 |
| ***Molnupiravir vs. Nirmatrelvir/r*** |  |  |  |  |
| IPWs | 0.91 (0.74, 1.07) | 1.00 (0.88, 1.11) | -0.09 (-0.29, 0.11) | 0.381 |
| Double Robust | 0.90 (0.70, 1.11) | 1.00 (0.88, 1.13) | -0.10 (-0.35, 0.15) | 0.324 |
| Regression adjustment | 0.90 (0.74, 1.07) | 1.00 (0.88, 1.12) | -0.10 (-0.31, 0.12) | 0.375 |
| Truncated |  |  | -0.34 (-0.61, -0.08) | 0.011 |
| ***Remdesivir vs. Nirmatrelvir/r*** |  |  |  |  |
| IPWs | 0.63 (0.51, 0.75) | 0.96 (0.84, 1.08) | -0.33 (-0.50, -0.16) | <.001 |
| Double Robust | 0.63 (0.50, 0.77) | 0.94 (0.81, 1.07) | -0.31 (-0.50, -0.12) | <.001 |
| Regression adjustment | 0.66 (0.52, 0.79) | 0.94 (0.80, 1.07) | -0.28 (-0.47, -0.09) | 0.004 |
| Truncated |  |  | -0.48 (-0.74, -0.23) | <.001 |
| ***Remdesivir vs. Sotrovimab*** |  |  |  |  |
| IPWs | 0.69 (0.54, 0.84) | 0.60 (0.51, 0.68) | 0.09 (-0.08, 0.26) | 0.290 |
| Double Robust | 0.68 (0.53, 0.83) | 0.60 (0.51, 0.69) | 0.08 (-0.10, 0.26) | 0.303 |
| Regression adjustment | 0.67 (0.53, 0.81) | 0.61 (0.52, 0.70) | 0.06 (-0.11, 0.23) | 0.478 |
| Truncated |  |  | 0.00 (-0.18, 0.18) | 0.985 |
| ***Sotrovimab vs. Nirmatrelvir/r*** |  |  |  |  |
| IPWs | 0.58 (0.50, 0.67) | 0.93 (0.79, 1.06) | -0.34 (-0.50, -0.18) | <.001 |
| Double Robust | 0.59 (0.49, 0.68) | 0.87 (0.65, 1.09) | -0.28 (-0.53, -0.04) | 0.017 |
| Regression adjustment | 0.59 (0.49, 0.69) | 0.93 (0.77, 1.09) | -0.34 (-0.53, -0.15) | <.001 |
| Truncated |  |  | -0.62 (-0.87, -0.38) | <.001 |
| ***Molnupiravir vs. Remdesivir*** |  |  |  |  |
| IPWs | 0.87 (0.74, 0.99) | 0.65 (0.53, 0.77) | 0.22 (0.04, 0.39) | 0.014 |
| Double Robust | 0.84 (0.64, 1.04) | 0.66 (0.53, 0.79) | 0.18 (-0.07, 0.43) | 0.066 |
| Regression adjustment | 0.87 (0.72, 1.02) | 0.66 (0.54, 0.79) | 0.20 (0.01, 0.40) | 0.044 |
| Truncated |  |  | 0.24 (0.00, 0.47) | 0.048 |
| ^&^Average Treatment Effect | | | | |
| ^*^weighted for month of infusion, duration of symptoms and immunodeficiency | | | | |

**Supplementary Table 4.** **Proportion of participants with undetectable Sars-CoV-2 in nasopharyngeal swab collected at day 7 (defined as cycle threshold, CT≤40) by intervention.**

|  | **Regimen started** | | | | | |
| --- | --- | --- | --- | --- | --- | --- |
|  | **Sotrovimab** | **Molnupiravir** | **Remdesivir** | **Nirmatrelvir/r** | **Total** | **p-value^*^** |
|  | **n (%)** | **n (%)** | **n (%)** | **n (%)** | **n (%)** |  |
| **Omicron (BA.1 + BA.2)** |  |  |  |  |  |  |
| Day7 CT > 40 | 8 (3.96) | 9 (7.69) | 7 (5.93) | 11 (13.10) | 35 (6.72) | 0.0001 |
| Day7 CT ≤ 40 | 194 (96.04) | 108 (92.31) | 111 (94.07) | 73 (86.90) | 486 (93.28) |  |
| **Omicron BA.1** |  |  |  |  |  |  |
| Day7 CT > 40 | 8 (5.48) | 9 (9.09) | 7 (8.33) | 7 (14.29) | 31 (8.20) | 0.0010 |
| Day7 CT ≤ 40 | 138 (94.52) | 90 (90.91) | 77 (91.67) | 42 (85.71) | 347 (91.80) |  |
| **Omicron BA.2** |  |  |  |  |  |  |
| Day7 CT > 40 | 0 (0) | 0 (0) | 0 (0) | 4 (11.43) | 4 (2.80) | 0.0031 |
| Day7 CT ≤ 40 | 56 (100) | 18 (100) | 34 (100) | 31(88.57) | 139 (97.20) |  |
| ^*^Fisher’s exact test | | | | | | |

**Supplementary figure 1: Variation of SARS-CoV-2 RNA levels from day1 to day7 in patients treated with Sotrovimab, Molnupiravir, Remdesivir, and Nirmatrelvir/r.**

Spaghetti-plots showing the log2 CT values measured at day1 (D1) and day7 (D7) in **(A)** each patient with

Omicron BA.1 infection treated with Sotrovimab (n=146), or Molnupiravir (n=99), or Remdesivir (n=84), or Nirmatrelvir/r (n=49); **(B)** in each patient with Omicron BA.2 infection treated with Sotrovimab (n=56), or Molnupiravir (n=18), or Remdesivir (n=34), or Nirmatrelvir/r (n=35)

Viral RNA levels are expressed as log2 CT values. CT values at D1 and D7 within each group were compared using paired Wilcoxon sign-rank test.

Horizontal dashed line represents the limit of detection (CT: 40.0); values≥40 are considered negative.

**Funding**

This study was supported by funds to the National institute for Infectious Diseases “*Lazzaro Spallanzani*”, IRCCS, Rome (Italy), from Italian Ministry of Health (Programme CCM 2020; Ricerca Corrente - Linea 1 on emerging and re-emerging infections) and from the European Commission - Horizon 2020 (CoNVat, Grant agreement ID 101003544; KRONO, Grant agreement ID 101005075).

**Declarations of interest:**

A.A. declares consultancy fees from Gilead Sciences, Merck, GSK, Pfizer, Astra Zeneca, and institutional research grants from Gilead Sciences and the Italian Medicine Agency (AIFA).

**Acknowledgments**

The authors gratefully acknowledge the nurse staff, all the patients, and all members of the **INMI COVID-19 Outpatient Treatment Study Group:**

Samir Al-Moghazi, Chiara Agrati, Andrea Antinori, Tommaso Ascoli-Bartoli, Francesco Baldini, Bartolini Barbara, Rita Bellagamba, Giulia Berno, Nazario Bevilacqua, Elisa Biliotti, Giulia Bonfiglio, Licia Bordi, Elena Boschiavo, Marta Camici, Emanuela Caraffa, Rita Casetti, Carlotta Cerva, Stefania Cicalini, Claudia Cimaglia, Francesca Colavita, Angela Corpolongo, Rita Corso, Alessandro Cozzi Lepri, Gianpiero D’Offizi, Federico De Zottis, Silvia Di Bari, Virginia Di Bari, Francesca Di Bella, Davide Roberto Donno, Angela D’Urso, Lavinia Fabeni, Massimo Francalancia, Marisa Fusto, Roberta Gagliardini, Paola Gallì, Francesca Gavaruzzi, Letizia Giancola, Giuseppina Giannico, Emanuela Giombini, Enrico Girardi, Giulia Gramigna, Elisabetta Grilli, Susanna Grisetti, Cesare Ernesto Maria Gruber, Giuseppina Iannicelli, Daniela Inzeo, Eleonora Lalle, Alessandra Lamonaca, Simone Lanini, Fabio Leotta, Raffaella Libertone, Laura Loiacono, Gaetano Maffongelli, Alessandra Marani, Andrea Mariano, Ilaria Mastrorosa, Giulia Matusali, Valentina Mazzotta, Silvia Meschi, Eugenia, Milozzi, Annalisa Mondi, Vanessa Mondillo, Emanuele Nicastri, Giovanna Onnelli, Sandrine Ottou, Claudia Palazzolo, Fabrizio Palmieri, Sara Pantanella, Jessica Paulicelli, Carmela Pinnetti, Pierluca Piselli, Maria Maddalena Plazzi, Alessandra Oliva, Alessia Rianda, Silvia Rosati, Martina Rueca, Alessandra Sacchi, Giuseppe Sberna, Laura Scorzolini, Fabrizio Taglietti, Giuseppina Tarabù, Francesca Trotti, Francesco Vaia, Alessandra Vergori, Serena Vita, Pietro Vittozzi.
